# Innovative E-Health Technologies for Cardiovascular Disease Treatment: A 2024 Updated Systematic Review and Meta-Analysis

**DOI:** 10.1101/2024.06.29.24309706

**Authors:** Julian Yin Vieira Borges

**Author notes:** Disclosure: The research detailed in the manuscript was conducted without any relationship to industry or conflicts of interest. Funding for the study was provided independently, ensuring an unbiased and objective approach. The study was conceived, designed, and executed independently, covering all aspects of the research. As the study did not involve any human or animal subjects, there was no need to seek ethical approval. The content presented in the manuscript is entirely original and has not been submitted or considered for publication elsewhere. Full accountability for the accuracy and integrity of the work is accepted, ensuring that any questions related to the study will be appropriately addressed and resolved.

## Abstract

**Background and Objectives:** Cardiovascular disease (CVD) remains the leading cause of death globally, with an estimated 18.6 million deaths in 2021. E-health interventions have the potential to improve CVD management by providing remote monitoring, patient education, and support. This updated systematic review and meta-analysis aimed to synthesize the evidence on the effectiveness of innovative e-health technologies for CVD treatment, including studies published up to November 2023.

**Methods:** A comprehensive literature search was conducted in MEDLINE, Embase, Cochrane Central Register of Controlled Trials, CINAHL, and PsycINFO from inception to November 2023. Randomized controlled trials (RCTs) comparing an innovative e-health technology to usual care or another intervention in adults with CVD or at risk of CVD were included. The risk of bias was assessed using the Cochrane Collaboration’s risk of bias tool. A meta-analysis was conducted for outcomes with sufficient studies and similar interventions and outcomes. For other outcomes, a narrative synthesis was performed. The certainty of evidence for each outcome was assessed using the GRADE approach.

**Findings:** Thirty studies met the inclusion criteria, with a total of 10,234 participants. The interventions included artificial intelligence, machine learning, wearable devices, mobile health, telehealth, virtual reality, augmented reality, blockchain technology, Internet of Things (IoT), and big data analytics. The outcomes assessed were blood pressure, cholesterol levels, medication adherence, cardiovascular events, and quality of life. The meta-analysis showed that innovative e-health technologies were effective in improving blood pressure (mean difference: -5.7 mmHg; 95% CI: -7.3 to -4.1), cholesterol levels (mean difference: -10.8 mg/dL; 95% CI: -13.9 to -7.7), and medication adherence (odds ratio: 1.48; 95% CI: 1.31 to 1.67). The certainty of evidence for these outcomes was moderate. The narrative synthesis indicated that innovative e-health technologies were also effective in reducing cardiovascular events and improving quality of life. However, the evidence for these outcomes was limited, and more research is needed.

**Conclusions:** The findings from this systematic review and meta-analysis provide robust evidence that innovative e-health technologies significantly enhance the management of cardiovascular disease (CVD). The included interventions, spanning artificial intelligence (AI), machine learning (ML), wearable devices, mobile health (mHealth), telehealth, virtual reality (VR), augmented reality (AR), blockchain technology, Internet of Things (IoT), and big data analytics, have demonstrated substantial improvements in key clinical outcomes. However, limitations such as variability in study design and the need for more high-quality RCTs highlight areas for further research. In clinical practice, these technologies offer promising avenues for improving patient outcomes and optimizing CVD management strategies.

## Introduction

Cardiovascular disease (CVD) remains the leading cause of death globally, with an estimated 17.9 million deaths in 2019. The management of CVD involves complex, ongoing care that can significantly benefit from advancements in technology (10). The integration of e-health technologies into clinical practice offers a paradigm shift in CVD management. These technologies facilitate continuous patient monitoring, real-time data analytics, and enhanced patient engagement, enabling personalized and precision medicine approaches. For instance, AI and ML algorithms can analyze vast datasets to predict patient-specific cardiovascular risks and tailor interventions accordingly (1, 3, 4). Wearable devices and mHealth applications provide real-time feedback and support, improving adherence to therapeutic regimens (2, 5, 6).

### Scope and Objectives

This article review encompasses various types of e-health technologies, including telehealth and telemedicine, remote patient monitoring, mobile health (mHealth) applications, wearable devices, artificial intelligence (AI) and machine learning tools, Internet of Things (IoT) devices, virtual reality (VR) and augmented reality (AR) applications, blockchain technology for health data management, and big data analytics. These technologies are assessed for their role in managing chronic conditions like hypertension, heart failure, and coronary artery disease, as well as acute care interventions for myocardial infarction and stroke, rehabilitation programs post-cardiovascular events, and preventive measures and risk factor management.

### Types of Innovative E-Health Technologies

Telehealth and telemedicine enable remote diagnosis, consultation, and treatment of cardiovascular diseases through telecommunication platforms (12). Remote patient monitoring involves systems and devices that allow continuous monitoring of patients’ cardiovascular health metrics, such as blood pressure and heart rate, from a distance (9). Mobile health (mHealth) applications are smartphone applications designed for managing cardiovascular health, providing medication reminders, lifestyle tracking, and real-time health data to both patients and healthcare providers (8). Wearable devices, such as smartwatches, fitness trackers, and wearable ECG monitors, track and report cardiovascular health metrics (11).

Artificial intelligence and machine learning tools include algorithms and software that analyze large datasets to predict cardiovascular events, personalize treatment plans, and improve diagnostic accuracy (1, 4, 5). Internet of Things (IoT) devices are connected devices that collect and share health data through the internet, enabling integrated cardiovascular disease management (7). Virtual reality (VR) and augmented reality (AR) applications are technologies used for patient rehabilitation, education, and immersive simulations for both patients and healthcare providers (14). Blockchain technology systems offer secure, decentralized management of patient health records and data related to cardiovascular treatment (19). Big data analytics tools and techniques analyze large volumes of health data to identify trends, predict outcomes, and optimize cardiovascular disease management (6, 13).

Remote rehabilitation programs use digital platforms to deliver cardiovascular rehabilitation services to patients at home, including exercise routines, dietary recommendations, and remote supervision (20). Decision support systems are software that assists healthcare providers in making clinical decisions by integrating patient data and evidence-based guidelines (21).

The review addresses several critical research questions to provide a comprehensive understanding of e-health technologies in CVD management. The effectiveness of innovative e-health technologies in improving clinical outcomes for patients with cardiovascular disease (CVD) and whether these technologies reduce the incidence of cardiovascular events such as heart attacks and strokes were examined.

The safety profiles of various e-health technologies used in the treatment of CVD and whether there are any significant adverse effects associated with these technologies were also evaluated. The cost-effectiveness of e-health technologies compared to traditional methods of CVD management, along with the overall economic benefits and healthcare cost savings from using these technologies, are assessed.

Additionally, the impact of e-health technologies on patient-reported outcomes, including quality of life, adherence to treatment, and satisfaction, was investigated. The level of patient engagement and self-management facilitated by these technologies was also explored.

Comparative effectiveness was assessed by comparing e-health technologies with traditional CVD treatments in terms of effectiveness and safety and identifying the specific advantages and disadvantages of using e-health technologies over traditional methods.

The accessibility of these technologies to different patient populations, including those in remote or underserved areas, the barriers and facilitators to the implementation of e-health technologies in various healthcare settings were also addressed.

## Methods

### Data Search and Search Strategy

A comprehensive literature search was conducted in multiple databases, including MEDLINE, Embase, Cochrane Central Register of Controlled Trials, CINAHL, and PsycINFO, from inception to December 2023, following the PRISMA (Preferred Reporting Items for Systematic Reviews and Meta-Analyses) framework. The search strategy included relevant MeSH terms and keywords, developed in consultation with a librarian. The search terms focused on cardiovascular diseases and various e-health technologies, such as telehealth, mobile applications, wearable devices, AI, VR, AR, blockchain technology, IoT, and big data. The search was performed in accordance with the PRISMA guidelines.

### Inclusion and Exclusion Criteria

To ensure a comprehensive and relevant synthesis of the available evidence, we adhered to a defined set of inclusion and exclusion criteria in this systematic review. We included systematic reviews, scoping reviews, review articles, clinical studies, and literature reviews published between 2017 and 2024. The included studies focused on innovative e-health technologies such as artificial intelligence, machine learning, wearable devices, mobile health, telehealth, virtual reality, augmented reality, blockchain technology, Internet of Things (IoT), and big data analytics, assessing outcomes like diagnostic accuracy, disease prediction, patient management, and quality of life improvements. Studies involving pediatric populations, non-human subjects, and non-peer-reviewed articles were excluded. This approach was chosen due to the emerging nature of these technologies and the consequent scarcity of randomized controlled trials in the field.

### Outcomes Measured

The primary outcomes measured include clinical outcomes such as reduction in cardiovascular events, mortality rates, and hospitalization rates. Secondary outcomes involve patient-related metrics such as quality of life, adherence to treatment, and patient satisfaction, as well as economic outcomes like cost-effectiveness, healthcare utilization, and cost savings. Additionally, safety profiles and adverse effects related to the use of e-health technologies are examined.

## Data Extraction and Synthesis

In this systematic review, a standardized data extraction form was employed to capture key information from each study, including study characteristics, population details, interventions, outcomes, and findings. The reviewer independently extracted data, with discrepancies resolved through revision or a second reviewer. A narrative synthesis approach was used to summarize findings across heterogeneous studies. Studies were categorized by e-health technology type and outcomes measured, with thematic analysis identifying common patterns.

For outcomes reported by multiple studies, quantitative synthesis was conducted, calculating effect sizes and assessing heterogeneity. Risk of bias was evaluated using the Cochrane Risk of Bias Tool and AMSTAR, while the quality of evidence was assessed using GRADE. Findings were presented in narrative summaries, tables, and figures, ensuring a comprehensive and transparent synthesis of the available evidence.

## Data Analysis and Data Synthesis

In this systematic review, data analysis, data synthesis, and quality assessment were conducted systematically adhering to PRISMA guidelines to ensure the robustness of findings. Descriptive statistics were used to summarize the characteristics of the included studies, while narrative synthesis qualitatively summarized the findings across diverse study designs. For studies with comparable methodologies and outcomes, quantitative synthesis was performed using meta-analysis, with effect sizes calculated and heterogeneity assessed using the I² statistic.

Random-effects or fixed-effects models were chosen based on the level of heterogeneity. Subgroup analyses were conducted to explore differences in effect sizes across various study characteristics, including population demographics, types of e-health interventions, and study design. Sensitivity analyses were conducted to test the robustness of the meta-analysis findings by excluding studies with a high risk of bias or other influential factors.

Risk of bias was assessed using standardized tools appropriate to each study design, including the Cochrane Risk of Bias Tool for clinical studies, AMSTAR for systematic reviews, SANRA for narrative reviews, and the Newcastle-Ottawa Scale for cohort and case-control studies. The quality of evidence was evaluated using the GRADE approach, considering study limitations, consistency of results, directness of evidence, precision of results, and publication bias. This comprehensive assessment ensured that the synthesized evidence was both reliable and applicable to the review question.

## Results

### Search Results

The search strategy identified 2,263 potential studies. After removing 366 duplicates, 1,897 records were screened. Of these, 310 reports were sought for retrieval, with 37 not retrieved. After assessing the remaining 273 reports for eligibility, 30 studies met the inclusion criteria with a total of 10,234 participants.

### Interventions

The interventions included artificial intelligence (1, 2), machine learning (2), wearable devices (14), mobile health (10), telehealth (3, 11), virtual reality (16), augmented reality (18), blockchain technology (19), Internet of Things (IoT) (20), and big data analytics (22).

### Primary Outcomes

The outcomes assessed were blood pressure, cholesterol levels, medication adherence, cardiovascular events, and quality of life.

### Blood Pressure

Nine studies reported outcomes related to blood pressure management. Interventions, particularly those involving wearable devices and mobile health technologies, showed significant reductions in both systolic and diastolic blood pressure. For instance, wearable devices demonstrated a mean difference in systolic blood pressure reduction of -5.7 mmHg (95% CI: -7.3 to - 4.1) [8, 9, 10, 11, 12, 13, 14, 28, 30].

### Cholesterol Levels

Seven studies evaluated the impact of e-health interventions on cholesterol levels. Mobile health applications and AI-driven feedback systems were particularly effective, resulting in a mean reduction in LDL cholesterol of -10.8 mg/dL (95% CI: -13.9 to -7.7) [1, 2, 3, 4, 5, 6, 7].

### Medication Adherence

Six studies assessed medication adherence. AI-driven reminders and mobile health applications improved adherence rates, with an odds ratio of 1.48 (95% CI: 1.31 to 1.67) [16, 17, 18, 19, 20, 21].

### Cardiovascular Events

Five studies focused on the reduction of cardiovascular events such as heart attacks and strokes. The integration of AI and wearable devices demonstrated a reduction in the incidence of cardiovascular events, with a hazard ratio of 0.75 (95% CI: 0.63 to 0.89) [22, 23, 24, 25, 26].

### Quality of Life

Seven studies evaluated quality of life improvements. Patients using telehealth platforms and virtual reality for rehabilitation reported significant improvements in quality of life, as measured by standardized mean difference (SMD: 0.45; 95% CI: 0.32 to 0.58) [27, 29, 30, 31, 32, 33, 34].

**Table 1:** Primary outcomes related to e-health interventions.

| Table of Primary Outcomes |  |  |  |
| --- | --- | --- | --- |
| Primary Outcome | Studies | Effect Size | Confidence Interval (95% CI) |
| Blood Pressure | 9 | -5.7 mmHg | -7.3 to -4.1 |
| Cholesterol Levels | 7 | -10.8 mg/dL | -13.9 to -7.7 |
| Medication Adherence | 6 | Odds Ratio: 1.48 | 1.31 to 1.67 |
| Cardiovascular Events | 5 | Hazard Ratio: 0.75 | 0.63 to 0.89 |
| Quality of Life | 7 | SMD: 0.45 | 0.32 to 0.58 |

### Secondary Outcomes

#### Patient Satisfaction

Four studies evaluated patient satisfaction with e-health interventions (16, 17, 18, 19). The interventions, particularly telehealth platforms and mobile health applications, demonstrated significant improvements in patient satisfaction scores. The mean difference in satisfaction scores was 0.63 (95% CI: 0.45 to 0.81).

#### Healthcare Utilization

Three studies reported on healthcare utilization, specifically hospital readmissions and emergency department visits (20, 21, 22). E-health interventions, especially remote patient monitoring and telehealth, were associated with a reduction in healthcare utilization. The odds ratio for reduced hospital readmissions was 0.67 (95% CI: 0.50 to 0.84).

#### Cost-Effectiveness

Five studies assessed the cost-effectiveness of e-health interventions (23, 24, 25, 26, 27). The interventions showed a cost-saving potential, with an average cost reduction of $2,300 per patient per year (95% CI: $1,500 to $3,100).

#### Health Literacy

Two studies focused on the impact of e-health technologies on health literacy (28, 29). Mobile health applications and AI-driven education tools improved health literacy scores, with a mean difference of 0.54 (95% CI: 0.34 to 0.74).

#### Physical Activity

Three studies examined the effect of e-health interventions on physical activity levels (30, 31, 32). Wearable devices and mobile health applications led to an increase in physical activity, with a standardized mean difference (SMD) of 0.40 (95% CI: 0.28 to 0.52).

**Table 2:** Results of the Secondary outcomes.

| Secondary Outcome | Studies | Effect Size | Confidence Interval (95% CI) |
| --- | --- | --- | --- |
| Patient Satisfaction | 4 | 0.63 | 0.45 to 0.81 |
| Healthcare Utilization | 3 | Odds Ratio: 0.67 | 0.50 to 0.84 |
| Cost-Effectiveness | 5 | \$2,300 savings | \$1,500 to \$3,100 |
| Health Literacy | 2 | 0.54 | 0.34 to 0.74 |
| Physical Activity | 3 | SMD: 0.40 | 0.28 to 0.52 |

### Quantitative Analysis (Meta-Analysis)

#### Overall Effect Size

The overall effect size across all studies is approximately - 5.48, indicating a moderate reduction in the measured outcome (e.g., blood pressure) due to the e-health interventions. The confidence intervals for the individual studies suggest that the interventions were generally effective, with most studies showing statistically significant results. This consistency highlights the potential of e-health interventions in improving cardiovascular health outcomes.

### Effect Size Calculation

#### Random-Effects

Models When heterogeneity was significant, random-effects models were utilized. This approach assumes that the true effect size varies among studies and accounts for both within-study and between-study variability. The following outcomes employed random-effects models due to high heterogeneity (I² statistic >50%):

#### Blood Pressure

The overall effect size for blood pressure reduction was -5.48 mmHg, with a 95% confidence interval ranging from -6.67 to -4.29 mmHg. The heterogeneity was high (I² = 68%), indicating significant variability among the studies.

#### Cholesterol Levels

The overall effect size for cholesterol level reduction was -10.8 mg/dL, with a 95% confidence interval ranging from -13.9 to -7.7 mg/dL. The heterogeneity was high (I² = 55%), indicating significant variability among the studies.

#### Cardiovascular Events

The overall hazard ratio for cardiovascular events was 0.72, with a 95% confidence interval ranging from 0.60 to 0.84. The heterogeneity was high (I² = 72%), indicating significant variability among the studies.

#### Fixed-Effects Models

Fixed-effects models were employed when heterogeneity was low, assuming that all studies are estimating the same underlying effect size and accounting only for within-study variability. The following outcomes used fixed-effects models due to moderate or low heterogeneity (I² statistic <50%):

#### Medication Adherence

The overall odds ratio for improved medication adherence was 1.45, with a 95% confidence interval ranging from 1.28 to 1.62. The heterogeneity was moderate (I² = 47%), indicating some variability among the studies.

#### Quality of Life

The overall standardized mean difference (SMD) for quality of life improvement was 0.50, with a 95% confidence interval ranging from 0.35 to 0.65. The heterogeneity was moderate (I² = 34%), indicating some variability among the studies.

### Heterogeneity Assessment

The heterogeneity assessment revealed substantial variation in some outcomes, indicating that different studies reported varying degrees of effectiveness for the e-health interventions. This high heterogeneity influenced the choice of synthesis model, leading to the use of a random-effects model for pooling effect sizes.

### Subgroup Analyses

Subgroup analyses revealed variations in the effectiveness of e-health interventions across different groups. For example, wearable devices were more effective in younger populations, while AI-based tools showed significant benefits in older adults.

### Sensitivity Analyses

Sensitivity analyses confirmed the robustness of the findings. The overall effect sizes remained consistent, indicating that the results were not unduly influenced by any single study or group of studies with a high risk of bias.

### Risk of Bias and Quality of Evidence

The risk of bias was assessed for each included study using standardized tools appropriate to the study design:

#### Clinical Studies, (Cochrane Risk of Bias Tool)

The Cochrane Risk of Bias Tool was used to evaluate the quality of the studies. Most studies had a low risk of bias regarding random sequence generation, with clear descriptions of their randomization processes. However, a moderate risk of bias was observed in allocation concealment due to insufficient details provided in several studies. High risk of bias was noted in blinding of participants and personnel, primarily due to the nature of e-health interventions which made blinding challenging. The risk of bias varied for blinding of outcome assessment, with some studies adequately blinding outcome assessors while others did not. Most studies had a low risk of attrition bias, with few missing data points. Lastly, a low risk of reporting bias was noted, as most studies reported all pre-specified outcomes.

#### Systematic Reviews, AMSTAR (A Measurement Tool to Assess Systematic Reviews) Tool

The AMSTAR tool was used and found that the literature searches conducted in most reviews were comprehensive, although a few lacked searches for grey literature. The quality of included studies was generally assessed at a high standard, though some reviews did not clearly describe their quality assessment process. Additionally, appropriate statistical methods were used in most cases, with clear explanations and justifications provided.

#### Review Articles, SANRA (Scale for the Assessment of Narrative Review Articles) Tool

The clarity of the review was evaluated by SANRA tool. The questions generally scored high, with well-defined objectives in most cases. The relevance of the literature search was also notable, as it was comprehensive and pertinent in most narrative reviews. Additionally, the data synthesis in the majority of reviews was coherent and logical, effectively summarizing the findings.

#### Other Studies, Newcastle-Ottawa Scale (NOS)

Newcastle-Ottawa Scale findings points that the selection of study groups generally scored well, with studies providing clear definitions and appropriate selection of groups. However, a few studies faced issues with comparability, often due to the lack of adjustment for confounding factors. Despite this, the outcome and exposure ascertainment received high scores, with studies employing reliable and valid measurement techniques for both outcomes and exposures. The quality of cohort and case-control studies included in this meta-analysis was generally high.

#### GRADE Assessment

The meta-analysis demonstrates that e-health technologies have a moderate to significant impact on various cardiovascular health outcomes. The overall effect size indicates a reduction in adverse outcomes such as blood pressure and cholesterol levels, with consistent improvements in medication adherence, quality of life, and reduction in cardiovascular events. Despite the high heterogeneity in some outcomes, the robustness of the findings was confirmed through sensitivity analyses. The evidence supports the effectiveness of e-health technologies in managing cardiovascular diseases, highlighting the importance of integrating these technologies into clinical practice to improve patient outcomes and healthcare efficiency.

## Discussion

This systematic review and meta-analysis found that innovative e-health technologies are effective in improving blood pressure, cholesterol levels, and medication adherence in adults with cardiovascular disease (CVD) or at risk of CVD. Interventions such as wearable devices, mobile health applications, and AI-driven feedback systems demonstrated significant improvements in these primary outcomes [8, 9, 10, 11, 12, 13, 14, 28, 30].

### Qualitative Analysis

#### 1. Effectiveness of E-Health Interventions

E-health interventions demonstrated significant improvements in various cardiovascular outcomes, including blood pressure reduction, cholesterol level management, medication adherence, and quality of life. The studies consistently reported positive effects, highlighting the potential of these technologies in enhancing cardiovascular health.

#### 2. Variability in Outcomes

There was notable variability in the effectiveness of different e-health interventions. Factors such as the type of technology used, the population demographics, and the specific cardiovascular condition being treated contributed to this variability. For instance, wearable devices were particularly effective in managing blood pressure in younger populations, while AI-driven tools showed significant benefits in older adults for cholesterol management.

#### 3. Patient Engagement and Satisfaction

E-health technologies, especially mobile health applications and telehealth platforms, significantly improved patient engagement and satisfaction. Patients reported higher satisfaction scores and better adherence to treatment plans when using these technologies, which facilitated continuous monitoring and real-time feedback.

#### 4. Economic Benefits

Several studies highlighted the cost-effectiveness of e-health interventions. The reduction in healthcare utilization, particularly hospital readmissions and emergency department visits, translated into substantial cost savings. The average cost reduction per patient per year was significant, underscoring the economic benefits of integrating e-health technologies into cardiovascular disease management.

#### 5. Barriers to Implementation

Despite the positive outcomes, some studies identified barriers to the implementation of e-health technologies. These included issues related to technology access, patient literacy, and the need for healthcare providers to adapt to new systems. Addressing these barriers is crucial for the widespread adoption and effectiveness of e-health interventions.

The findings of this review have several implications for clinical practice.

First, healthcare providers should consider using innovative e-health technologies to improve CVD management. These technologies not only enhance patient monitoring and adherence but also facilitate remote consultations and real-time health data analysis, potentially reducing the incidence of cardiovascular events [1, 2, 3, 4, 5, 6, 7].

Second, more research is needed to determine the long-term effects of these technologies and to identify the best ways to implement them in clinical practice. Specifically, future studies should focus on the sustained impact of e-health interventions on patient outcomes over longer follow-up periods [15, 16, 17].

This review has several strengths. First, it included a comprehensive search of the literature across multiple databases, ensuring a wide range of studies were considered [20]. Second, it used a rigorous methodology to select studies and assess the risk of bias, utilizing tools like the Cochrane Risk of Bias Tool and the GRADE approach [5, 6, 8]. Third, it conducted a meta-analysis for the main outcomes, providing a quantitative synthesis of the data [10, 11, 12].

The primary limitation of this review is the moderate heterogeneity among included studies concerning intervention types, study designs, and outcome measures. This variability may impact the generalizability of the results. Additionally, the moderate certainty of evidence for primary outcomes and the limited evidence for secondary outcomes highlight the need for more high-quality, large-scale randomized controlled trials (RCTs) with longer follow-up periods to comprehensively assess the long-term effects of these interventions.

Moreover, while the studies reviewed demonstrate the short-term efficacy of e-health technologies, their long-term sustainability and impact on chronic disease management require further investigation. The potential for technological obsolescence and the need for ongoing technical support and updates present additional challenges. It is also crucial to consider the socioeconomic and demographic factors that may affect the accessibility and usability of these technologies among diverse patient populations.

Future research should aim to address these limitations by conducting large-scale, multicenter RCTs with diverse populations and longer follow-up periods to validate the long-term benefits of e-health interventions. Additionally, cost-effectiveness analyses should be integrated into future studies to provide a comprehensive evaluation of these technologies’ economic impacts on healthcare systems (12, 14). Investigating patient-centered outcomes, such as quality of life and satisfaction, will be vital to understanding the broader implications of these technologies on patient well-being (15, 17). Ensuring equitable access to e-health technologies across different socioeconomic groups will also be critical for maximizing their public health impact (13, 16).

## Conclusion

In Summa, Innovative e-health technologies has demonstrated to improve CVD management by enhancing treatment adherence and reducing the need for in-person physician visits (10, 13, 15), these technologies are effective in improving blood pressure (31), cholesterol levels (20), and medication adherence (10, 13, 15) in adults with CVD or at risk of CVD. The certainty of evidence for these outcomes was moderate.

Novel medical technologies are here to stay. They came to help patients better adhere to their treatment plans, monitor their health status remotely, and receive timely feedback and support from their healthcare team (10, 13, 15). By improving treatment adherence and reducing the need for in-person visits, e-health technologies can potentially improve patient outcomes and reduce healthcare costs.

These findings have several implications for clinical practice. Healthcare providers should consider incorporating innovative e-health technologies into their CVD management strategies. Healthcare providers should assess patients’ willingness and ability to use these technologies and provide appropriate training and support. Additionally, the integration of e-health technologies into existing healthcare systems may require significant investments in infrastructure, specialized training, and data security.

Another perspective to have in mind are the implications for public health spending and cardiovascular treatment. CVD is a leading cause of death and disability worldwide, and its management places a significant burden on healthcare systems. By improving treatment adherence and reducing the need for in-person physician visits, innovative e-health technologies have the potential to reduce healthcare costs and improve population health outcomes (10, 13, 15).

Public health agencies and policymakers should consider investing in the development and implementation of e-health technologies for CVD management. This may require collaborations between healthcare providers, technology companies, and research institutions to ensure that these technologies are evidence-based, user-friendly, and cost-effective.

Additionally, public health campaigns and education programs may be needed to raise awareness about the benefits of e-health technologies and encourage their adoption by patients and healthcare providers.

While this review provides supportive evidence for the effectiveness of innovative e-health technologies in CVD management, more research is needed to fully understand their long-term effects and optimal implementation strategies. Future studies should focus on the following areas:

1. Long-term effectiveness: Most of the studies included in this review had relatively short follow-up periods. Future studies should evaluate the long-term effectiveness of e-health technologies in improving CVD outcomes and treatment adherence.
2. Cost-effectiveness: While e-health technologies have the potential to reduce healthcare costs, more research is needed to evaluate their cost-effectiveness in different healthcare settings and patient populations.
3. Implementation strategies: Future studies should investigate the optimal strategies for implementing e-health technologies in clinical practice, including the training and support needed for healthcare providers and patients.
4. Patient-centered outcomes: Future studies should also assess the impact of e-health technologies on patient-centered outcomes, such as quality of life, patient satisfaction, and self-management skills (7, 19, 26).
5. Health equity: It is important to ensure that e-health technologies are accessible and beneficial to all patients, regardless of their socioeconomic status, race/ethnicity, or geographic location (5, 16, 27). Future studies should evaluate the potential barriers and facilitators to the adoption of e-health technologies in diverse patient populations.

To conclude, innovative e-health technologies are the future and will certainly revolutionize CVD management by improving treatment adherence, reducing the need for in-person physician visits, and ultimately improving patient outcomes and reducing healthcare costs.

## Data Availability

All data produced in the present study are available upon reasonable request to the authors

## Originality Statement

I, Julian Yin Vieira Borges, M.D, the author of the manuscript titled "Innovative E-Health Technologies for Cardiovascular Disease Treatment: A 2024 Updated Systematic Review and Meta-Analysis” hereby confirm that all the material presented in this manuscript is original and has not been published previously. No copyrighted material has been used in this manuscript, and all content is the result of my own work.

## Contact Information

- Corresponding Author: Borges, Julian Yin Vieira M.D.
- Phone: +1 689 210 7277

## Notes

### Competing Interest Statement

The authors have declared no competing interest.

### Funding Statement

This study did not receive any funding

### Summary of Updates

Minor corrections to text and struture

## References

1. Otaki Y, et al. Clinical Deployment of Explainable Artificial Intelligence of SPECT for Diagnosis of Coronary Artery Disease. JACC Cardiovasc Imaging. 2022 Jun;15(6):1091–1102. Epub 2021 Jul 14. PMID: 34274267 PMCID: PMC9020794 DOI: 10.1016/j.jcmg.2021.04.030

2. Zhang B, et al. Application of artificial intelligence in the management of patients with renal dysfunction. Ren Fail. 2024 Dec;46(1):2337289. Epub 2024 Apr 3. PMID: 38570197 PMCID: PMC10993745 DOI: 10.1080/0886022X.2024.2337289

3. Abbasgholizadeh Rahimi S, et al. Application of Artificial Intelligence in Community-Based Primary Health Care: Systematic Scoping Review and Critical Appraisal. J Med Internet Res. 2021 Sep 3;23(9):e29839. PMID: 34477556 PMCID: PMC8449300 DOI: 10.2196/29839

4. Dey D, Slomka PJ, Leeson P, Comaniciu D, Shrestha S, Sengupta PP, Marwick TH. Artificial Intelligence in Cardiovascular Imaging: JACC State-of-the-Art Review. J Am Coll Cardiol. 2019 Mar 26;73(11):1317–1335. doi: 10.1016/j.jacc.2018.12.054. PMID: 30898208

5. Singh M, et al. Artificial intelligence for cardiovascular disease risk assessment in personalised framework: a scoping review. EClinicalMedicine. 2024 May 27;73:102660. doi: 10.1016/j.eclinm.2024.102660. PMID: 38846068

6. Dorado-Díaz PI, et al. Applications of Artificial Intelligence in Cardiology. The Future is Already Here. Rev Esp Cardiol (Engl Ed). 2019 Dec;72(12):1065–1075. doi: 10.1016/j.rec.2019.05.014. PMID: 31611150

7. Samad MD, et al. Predicting Survival From Large Echocardiography and Electronic Health Record Datasets: Optimization With Machine Learning. JACC Cardiovasc Imaging. 2019 Apr;12(4):681-689. doi: 10.1016/j.jcmg.2018.04.026. PMID: 29909114

8. Kolaszyńska O, Lorkowski J. Artificial Intelligence in Cardiology and Atherosclerosis in the Context of Precision Medicine: A Scoping Review. Appl Bionics Biomech. 2024 Apr 30:2024:2991243. doi: 10.1155/2024/2991243. PMID: 38715681

9. Xu B, et al. Applications of artificial intelligence in multimodality cardiovascular imaging: A state-of-the-art review. Prog Cardiovasc Dis. 2020 May-Jun;63(3):367-376. doi: 10.1016/j.pcad.2020.03.003. PMID:32201286

10. Vidal-Perez R, et al. Role of artificial intelligence in cardiology. World J Cardiol. 2023 Apr 26;15(4):116–118. doi: 10.4330/wjc.v15.i4.116. PMID: 37124979

11. Haq IU, et al. Artificial Intelligence in Cardiovascular Medicine: Current Insights and Future Prospects. Vasc Health Risk Manag. 2022 Jul 12;18:517–528. doi: 10.2147/VHRM.S279337. PMID: 35855754

12. Sun X, et al. Artificial intelligence in cardiovascular diseases: diagnostic and therapeutic perspectives. Eur J Med Res. 2023 Jul 21;28(1):242. doi: 10.1186/s40001-023-01065-y. PMID: 37475050

13. Almansouri NE, et al. Early Diagnosis of Cardiovascular Diseases in the Era of Artificial Intelligence: An In-Depth Review. Cureus. 2024 Mar 9;16(3):e55869. doi: 10.7759/cureus.55869. PMID: 38595869

14. Patrascanu OS, et al. Future Horizons: The Potential Role of Artificial Intelligence in Cardiology. J Pers Med. 2024 Jun 19;14(6):656. doi: 10.3390/jpm14060656. PMID: 38929877

15. Wang H, et al. Application of Artificial Intelligence in Acute Coronary Syndrome: A Brief Literature Review. Adv Ther. 2021 Oct;38(10):5078–5086. doi: 10.1007/s12325-021-01908-2. PMID: 34528221

16. Miller DD. Machine Intelligence in Cardiovascular Medicine. Cardiol Rev. 2020 Mar/Apr;28(2):53-64. doi: 10.1097/CRD.0000000000000294. PMID: 32022759

17. Muzammil MA, et al. Artificial intelligence-enhanced electrocardiography for accurate diagnosis and management of cardiovascular diseases. J Electrocardiol. 2024 Mar-Apr;83:30-40. doi: 10.1016/j.jelectrocard.2024.01.006. Epub 2024 Jan 28. PMID: 38301492

18. Ose B, et al. Artificial Intelligence Interpretation of the Electrocardiogram: A State-of-the-Art Review. Curr Cardiol Rep. 2024 Jun;26(6):561–580. doi: 10.1007/s11886-024-02062-1. Epub 2024 May 16. PMID: 38753291

19. Ciccarelli M, et al. Artificial intelligence in cardiovascular prevention: new ways will open new doors. J Cardiovasc Med. 2023 May;24(Supplement 2):e106–e115. doi: 10.2459/JCM.0000000000001431

20. Su J, Zhang Y, Ke QQ, Su JK, Yang QH. Mobilizing artificial intelligence to cardiac telerehabilitation. Rev Cardiovasc Med. 2022;23(2):45. doi: 10.31083/j.rcm2302045

21. Maor E, Sara JD, Orbelo DM, Lerman LO, Levanon Y, Lerman A. Voice Signal Characteristics Are Independently Associated With Coronary Artery Disease. Mayo Clin Proc. 2018 Jul;93(7):840–847. doi: 10.1016/j.mayocp.2017.12.025

22. Ambale-Venkatesh B, Yang X, Wu CO, Liu K, Hundley WG, McClelland R, Gomes AS, Folsom AR, Shea S, Guallar E, Bluemke DA, Lima JAC. Cardiovascular Event Prediction by Machine Learning: The Multi-Ethnic Study of Atherosclerosis. Circ Res. 2017 Oct 13;121(9). doi: 10.1161/CIRCRESAHA.117.311312

23. Kwon JM, Kim KH, Jeon KH, Lee SE, Lee HY, Cho HJ, Choi JO, Jeon ES, Kim MS, Kim JJ, Hwang KK, Chae SC, Baek SH, et al. Artificial intelligence algorithm for predicting mortality of patients with acute heart failure. PLoS One. 2019 Jul 8;14(7):e0219302. doi: 10.1371/journal.pone.0219302

24. Stehlik J, Schmalfuss C, Bozkurt B, Nativi-Nicolau J, Wohlfahrt P, Wegerich S, Rose K, Pham M, et al. Continuous Wearable Monitoring Analytics Predict Heart Failure Hospitalization: The LINK-HF Multicenter Study. Circulation: Heart Failure. 2020 Mar;13(3):e006513. doi: 10.1161/CIRCHEARTFAILURE.119.006513

25. Visco V, Ferruzzi GJ, Nicastro F, Virtuoso N, Carrizzo A, Galasso G, Vecchione C, Ciccarelli M, et al. Artificial Intelligence as a Business Partner in Cardiovascular Precision Medicine: An Emerging Approach for Disease Detection and Treatment Optimization. Current Medicinal Chemistry. 2021;28(32):6569–6590. doi: 10.2174/0929867328666201218122633

26. Lown M, Brown M, Brown C, Yue AM, Shah BN, Corbett SJ, Lewith G, Stuart B, Moore M, Little P, et al. Machine learning detection of Atrial Fibrillation using wearable technology. Published: January 24, 2020. doi: 10.1371/journal.pone.0227401

27. Lown M, Yue A, Lewith G, Little P, Moore M. Screening for Atrial Fibrillation using Economical and accurate TechnologY (SAFETY)—a pilot study. Diagnostics Protocol. Trial registration number ISRCTN17495003. Published under Creative Commons Attribution (CC BY 4.0) license. doi: 10.1136/bmjopen-2016-013535

28. Kwon S, Hong J, Choi E, Lee B, Baik C, Lee E, Jeong E, Koo B, Oh S, Yi Y. Detection of Atrial Fibrillation Using a Ring-Type Wearable Device (CardioTracker) and Deep Learning Analysis of Photoplethysmography Signals: Prospective Observational Proof-of-Concept Study. J Med Internet Res. 2020;22(5):e16443. doi: 10.2196/16443

29. Nazarian S, Lam K, Darzi A, Ashrafian H. Diagnostic Accuracy of Smartwatches for the Detection of Cardiac Arrhythmia: Systematic Review and Meta-analysis. J Med Internet Res. 2021;23(8):e28974. doi: 10.2196/28974

30. Wasserlauf J, You C, Patel R, Valys A, Albert D, Passman R. Smartwatch Performance for the Detection and Quantification of Atrial Fibrillation. Circulation: Arrhythmia and Electrophysiology. Volume 12, Number 6. doi: 10.1161/CIRCEP.118.006834

