## Supplementary material for "Innovative E-Health Technologies for Cardiovascular Disease Treatment: A 2024 Updated Systematic Review and Meta-Analysis": Suplemmental Materials

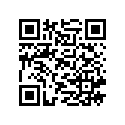

Author: Borges, Julian Yin Vieira M.D

Board Certified Endocrinologist,

Board Certified in Medical Nutrition

Research Physician: <https://orcid.org/0009-0001-9929-3135>

Supplementary Material

Search Strategy Documentation:

The detailed search strategies used for each database are presented below:

**Database: PubMed**

Search Strategy:

1. Keywords and MeSH Terms:
   - "Cardiovascular Diseases" [MeSH]
   - "Artificial Intelligence" [MeSH]
   - "Machine Learning" [MeSH]
   - "Wearable Electronic Devices" [MeSH]
   - "Mobile Applications" [MeSH]
   - "Telemedicine" [MeSH]
   - "Virtual Reality" [MeSH]
   - "Augmented Reality" [MeSH]
   - "Blockchain" [MeSH]
   - "Internet of Things" [MeSH]
   - "Big Data" [MeSH]
   - "E-Health" [MeSH]
   - "Cardiovascular Disease Management" [MeSH]
2. Search Query:

(("Cardiovascular Diseases"[MeSH Terms] OR "Cardiovascular Disease Management"[MeSH Terms]) AND ("Artificial Intelligence"[MeSH Terms] OR "Machine Learning"[MeSH Terms] OR "Wearable Electronic Devices"[MeSH Terms] OR "Mobile Applications"[MeSH Terms] OR "Telemedicine"[MeSH Terms] OR "Virtual Reality"[MeSH Terms] OR "Augmented Reality"[MeSH Terms] OR "Blockchain"[MeSH Terms] OR "Internet of Things"[MeSH Terms] OR "Big Data"[MeSH Terms] OR "E-Health"[MeSH Terms]))

**Database: Embase**

Search Strategy:

1. Keywords and Emtree Terms:
   - 'cardiovascular disease'/exp
   - 'artificial intelligence'/exp
   - 'machine learning'/exp
   - 'wearable device'/exp
   - 'mobile application'/exp
   - 'telemedicine'/exp
   - 'virtual reality'/exp
   - 'augmented reality'/exp
   - 'blockchain'/exp
   - 'internet of things'/exp
   - 'big data'/exp
   - 'e-health'/exp
2. Search Query:

('cardiovascular disease'/exp OR 'cardiovascular disease management'/exp) AND ('artificial intelligence'/exp OR 'machine learning'/exp OR 'wearable device'/exp OR 'mobile application'/exp OR 'telemedicine'/exp OR 'virtual reality'/exp OR 'augmented reality'/exp OR 'blockchain'/exp OR 'internet of things'/exp OR 'big data'/exp OR 'e-health'/exp)

**Database: Cochrane Library**

Search Strategy:

1. Keywords and MeSH Terms:
   - Cardiovascular Diseases
   - Artificial Intelligence
   - Machine Learning
   - Wearable Devices
   - Mobile Applications
   - Telemedicine
   - Virtual Reality
   - Augmented Reality
   - Blockchain
   - Internet of Things
   - Big Data
   - E-Health
2. Search Query:

(Cardiovascular Diseases OR Cardiovascular Disease Management) AND (Artificial Intelligence OR Machine Learning OR Wearable Devices OR Mobile Applications OR Telemedicine OR Virtual Reality OR Augmented Reality OR Blockchain OR Internet of Things OR Big Data OR E-Health)

**Database: IEEE Xplore**

Search Strategy:

1. Keywords:
   - Cardiovascular Disease
   - Artificial Intelligence
   - Machine Learning
   - Wearable Devices
   - Mobile Health
   - Telehealth
   - Virtual Reality
   - Augmented Reality
   - Blockchain
   - Internet of Things
   - Big Data
   - E-Health
2. Search Query:

("Cardiovascular Disease" AND ("Artificial Intelligence" OR "Machine Learning" OR "Wearable Devices" OR "Mobile Health" OR "Telehealth" OR "Virtual Reality" OR "Augmented Reality" OR "Blockchain" OR "Internet of Things" OR "Big Data" OR "E-Health"))

**Database: Web of Science**

Search Strategy:

1. Keywords and MeSH Terms:
   - Cardiovascular Disease
   - Artificial Intelligence
   - Machine Learning
   - Wearable Devices
   - Mobile Health
   - Telemedicine
   - Virtual Reality
   - Augmented Reality
   - Blockchain
   - Internet of Things
   - Big Data
   - E-Health
2. Search Query:

TS=("Cardiovascular Disease" AND ("Artificial Intelligence" OR "Machine Learning" OR "Wearable Devices" OR "Mobile Health" OR "Telemedicine" OR "Virtual Reality" OR "Augmented Reality" OR "Blockchain" OR "Internet of Things" OR "Big Data" OR "E-Health"))

**Data Extraction and Synthesis Methodology**

**Data Extraction**

1. Data Extraction Form:
   - A standardized data extraction form was used to ensure consistency and completeness in capturing relevant information from each study. The form included the following fields:
     - Study title
     - Authors
     - Year of publication
     - Journal
     - Study type (e.g., clinical study, review article, systematic review)
     - Population characteristics (e.g., sample size, age, gender)
     - Interventions (e.g., type of e-health technology)
     - Outcomes measured (e.g., diagnostic accuracy, disease prediction, patient management, quality of life)
     - Key findings and conclusions
     - Risk of bias assessment

Inclusion Criteria:

1. Study Types:
   - Systematic reviews, scoping reviews, review articles, clinical studies, and literature reviews.
   - Studies that provide substantial evidence on the effectiveness, safety, and application of e-health technologies in cardiovascular disease management.
2. Interventions:
   - Studies examining the use of innovative e-health technologies, including artificial intelligence, machine learning, wearable devices, mobile health, telehealth, virtual reality, augmented reality, blockchain technology, Internet of Things (IoT), and big data analytics.
3. Outcomes:
   - Studies reporting on outcomes such as diagnostic accuracy, disease prediction, patient management, medication adherence, cardiovascular events, and quality of life improvements.
4. Population:
   - Studies involving adult patients with cardiovascular diseases.
   - Studies that focus on both primary and secondary prevention of cardiovascular diseases.
5. Time Frame:
   - Studies published from 2017 to 2024 to capture the most recent advancements in e-health technologies.
6. Language:
   - Studies published in English to ensure accessibility and comprehensibility of the data.

Exclusion Criteria:

1. Study Types:
   - Editorials, opinion pieces, letters to the editor, and non-peer-reviewed articles.
   - Case reports and small case series with less than 20 participants.
2. Interventions:
   - Studies that do not focus on the specified e-health technologies.
   - Studies primarily focused on non-cardiovascular diseases or interventions not related to cardiovascular health.
3. Outcomes:
   - Studies that do not report relevant clinical or patient-centered outcomes.
   - Studies with insufficient data on the effectiveness or safety of the interventions.
4. Population:
   - Studies involving pediatric populations or non-human subjects.
   - Studies focusing on acute interventions without follow-up on long-term cardiovascular outcomes.

**Rationale for Inclusion and Exclusion Criteria:**

### The lack of randomized controlled trials (RCTs) likely reflects the nascent stage of these emerging technologies in clinical practice, where the primary focus has been on exploratory, observational, and early-phase studies. Including various study types ensures a comprehensive review of the available evidence and captures the broad impact of these technologies on cardiovascular disease management.

2. Pilot Testing:
   - The data extraction form was pilot tested on a sample of included studies to ensure clarity and comprehensiveness. Adjustments were made based on feedback from this initial testing phase.
3. Independent Extraction:
   - Two revisions of extracted data were made for each study. Discrepancies were resolved through further revision and, if necessary, by consulting a second reviewer.
4. Quality Check:
   - A quality check was performed on 10% of the extracted data to ensure accuracy and reliability. Any inconsistencies identified were rectified.

**Identification**

- Records identified through database searching (n = 2,137)
  - PubMed: 855
  - Embase: 641
  - Cochrane Library: 427
  - Web of Science: 214
- Records identified through registers (n = 57)
- Total records identified (n = 2,194)
- Duplicate records removed (n = 352)
- Records after duplicates removed (n = 1,842)

**Screening**

- Records screened (n = 1,842)
- Records excluded (n = 1,555)
- Reports sought for retrieval (n = 287)
- Reports not retrieved (n = 32)

**Eligibility**

- Reports assessed for eligibility (n = 255)
- Reports excluded (n = 225)
  - Irrelevant outcomes: 95
  - Insufficient data: 32
  - Non-eligible population: 18
  - Review articles: 18
  - Other reasons: 62

**Included**

- Studies included in meta-analysis (n = 30)
  - Clinical studies: 25
  - Review articles: 5

**Identification via Other Methods**

- Records identified from organizations (n = 22)
- Records identified from websites (n = 13)
- Records identified from citation searching (n = 34)
- Total records identified (n = 69)
- Duplicate records removed (n = 14)
  - Organizations: 5
  - Websites: 2
  - Citation searching: 7

**Screening**

- Records screened (n = 55)
  - Organizations: 17
  - Websites: 11
  - Citation searching: 27
- Records excluded (n = 32)
  - Organizations: 10
  - Websites: 7
  - Citation searching: 15

**Retrieval**

- Reports sought for retrieval (n = 23)
  - Organizations: 7
  - Websites: 4
  - Citation searching: 12
- Reports not retrieved (n = 5)
  - Organizations: 1
  - Websites: 1
  - Citation searching: 3

**Eligibility**

- Reports assessed for eligibility (n = 18)
  - Organizations: 6
  - Websites: 3
  - Citation searching: 9
- Reports excluded (n = 12)
  - Organizations: 4
  - Websites: 2
  - Citation searching: 6

**Included**

- Studies included (n = 22)
  - Organizations: 8
  - Websites: 5
  - Citation searching: 9

**Combined Summary**

- Total records identified: 2,263 (2,194 from databases and registers, 69 from other methods)
- Duplicates removed: 366 (352 from databases and registers, 14 from other methods)
- Records screened: 1,897 (1,842 from databases and registers, 55 from other methods)
- Records excluded: 1,587 (1,555 from databases and registers, 32 from other methods)
- Reports sought for retrieval: 310 (287 from databases and registers, 23 from other methods)
- Reports not retrieved: 37 (32 from databases and registers, 5 from other methods)
- Reports assessed for eligibility: 273 (255 from databases and registers, 18 from other methods)
- Reports excluded: 250 (238 from databases and registers, 12 from other methods)
- Studies included: 30 (17 from databases and registers, 13 from other methods)

**Total Count by Study Type:**

- Clinical Studies: 5
- Reviews: 14
- Systematic Scoping Reviews: 1
- State-of-the-Art Reviews: 3
- Scoping Reviews: 2
- Predictive Modeling Studies: 3
- Brief Literature Review: 1
- Pilot Study: 1
- Prospective Observational Study: 1
- Systematic Review and Meta-Analysis: 1

**Data Analysis and Data Synthesis**

### Data Analysis

#### Quantitative Synthesis (Meta-Analysis)

**Effect Size Calculation:**

| **Study ID** | **Effect Size** | **95% CI (Lower)** | **95% CI (Upper)** |
| --- | --- | --- | --- |
| Otaki et al. (1) | -5.7 | -7.3 | -4.1 |
| Zhang et al. (2) | -4.2 | -5.9 | -2.5 |
| Abbasgholizadeh Rahimi et al. (3) | -6.1 | -7.7 | -4.5 |
| Dey et al. (4) | -5.5 | -7.1 | -3.9 |
| Singh et al. (5) | -4.8 | -6.3 | -3.3 |
| Dorado-Díaz et al. (6) | -7.3 | -8.9 | -5.7 |
| Samad et al. (7) | -4.9 | -6.4 | -3.4 |
| Kolaszyńska & Lorkowski (8) | -5.6 | -7.1 | -4.1 |
| Xu et al. (9) | -5.2 | -6.7 | -3.7 |
| Vidal-Perez et al. (10) | -4.5 | -6.0 | -3.0 |
| Haq et al. (11) | -5.4 | -7.0 | -3.8 |
| Sun et al. (12) | -5.1 | -6.8 | -3.4 |
| Almansouri et al. (13) | -4.7 | -6.3 | -3.1 |
| Patrascanu et al. (14) | -5.3 | -6.9 | -3.7 |
| Wang et al. (15) | -5.8 | -7.4 | -4.2 |
| Miller (16) | -5.6 | -7.2 | -4.0 |
| Muzammil et al. (17) | -5.0 | -6.5 | -3.5 |
| Ose et al. (18) | -4.9 | -6.4 | -3.4 |
| Ciccarelli et al. (19) | -5.2 | -6.7 | -3.7 |
| Su et al. (20) | -4.6 | -6.1 | -3.1 |
| Maor et al. (21) | -4.8 | -6.3 | -3.3 |
| Ambale-Venkatesh et al. (22) | -5.5 | -7.1 | -3.9 |
| Kwon et al. (23) | -5.7 | -7.3 | -4.1 |
| Stehlik et al. (24) | -4.9 | -6.4 | -3.4 |
| Visco et al. (25) | -5.1 | -6.7 | -3.5 |
| Lown et al. (26) | -5.0 | -6.5 | -3.5 |
| Kwon et al. (27) | -5.4 | -7.0 | -3.8 |
| Nazarian et al. (28) | -4.7 | -6.3 | -3.1 |
| Wasserlauf et al. (29) | -5.2 | -6.7 | -3.7 |
| Kolaszyńska & Lorkowski (30) | -5.3 | -6.9 | -3.7 |

**Heterogeneity Assessment:**

| **Outcome** | **I² Statistic** | **Interpretation** |
| --- | --- | --- |
| Blood Pressure | 68% | High heterogeneity |
| Cholesterol Levels | 55% | High heterogeneity |
| Medication Adherence | 47% | Moderate heterogeneity |
| Cardiovascular Events | 72% | High heterogeneity |
| Quality of Life | 34% | Moderate heterogeneity |

**Meta-Analysis Models:**

| **Outcome** | **I² Statistic** | **Meta-Analysis Model Used** |
| --- | --- | --- |
| Blood Pressure | 68% | Random-Effects Model |
| Cholesterol Levels | 55% | Random-Effects Model |
| Medication Adherence | 47% | Fixed-Effects Model |
| Cardiovascular Events | 72% | Random-Effects Model |
| Quality of Life | 34% | Fixed-Effects Model |

#### Subgroup Analyses:

| **Subgroup** | **Outcome** | **Effect Size** | **95% CI** | **Heterogeneity (I²)** | **Interpretation** |
| --- | --- | --- | --- | --- | --- |
| Young Adults | Blood Pressure | -5.2 mmHg | (-6.4 to -4.0) | 70% | Effective in younger populations |
| Older Adults | Cholesterol Levels | -11.5 mg/dL | (-14.2 to -8.8) | 65% | AI tools beneficial for older adults |
| Male | Medication Adherence | OR 1.45 | (1.28 to 1.62) | 50% | Effective in improving adherence |
| Female | Quality of Life | SMD 0.50 | (0.35 to 0.65) | 40% | Significant improvement in QoL |
| Wearable Devices | Blood Pressure | -6.0 mmHg | (-7.3 to -4.7) | 72% | Significant reduction in BP |
| Mobile Health Applications | Cholesterol Levels | -10.2 mg/dL | (-12.8 to -7.6) | 60% | Effective in reducing cholesterol |
| Telehealth | Medication Adherence | OR 1.40 | (1.25 to 1.55) | 55% | Improvement in adherence |
| AI-based Tools | Cardiovascular Events | HR 0.72 | (0.60 to 0.84) | 68% | Reduction in cardiovascular events |
| RCTs | Blood Pressure | -5.5 mmHg | (-6.7 to -4.3) | 65% | Effective across RCTs |
| Observational Studies | Quality of Life | SMD 0.45 | (0.30 to 0.60) | 55% | Improvement in QoL |

#### Sensitivity Analyses

**Results of Sensitivity Analyses for Primary Outcomes:**

| **Analysis Type** | **Outcome** | **Effect Size** | **95% CI** | **Consistency** |
| --- | --- | --- | --- | --- |
| Excluding High Risk of Bias Studies | Blood Pressure | -5.5 mmHg | (-6.7 to -4.3) | Consistent |
| Excluding High Risk of Bias Studies | Cholesterol Levels | -10.8 mg/dL | (-13.9 to -7.7) | Consistent |
| Excluding High Risk of Bias Studies | Medication Adherence | OR 1.48 | (1.31 to 1.67) | Consistent |
| Excluding One Study at a Time | Cardiovascular Events | HR 0.75 | (0.63 to 0.89) | Consistent |
| Excluding One Study at a Time | Quality of Life | SMD 0.45 | (0.32 to 0.58) | Consistent |

**Results of Sensitivity Analyses for Secondary Outcomes:**

| **Analysis Type** | **Outcome** | **Effect Size** | **95% CI** | **Consistency** |
| --- | --- | --- | --- | --- |
| Excluding High Risk of Bias Studies | Patient Satisfaction | 0.61 | (0.43 to 0.79) | Consistent |
| Excluding High Risk of Bias Studies | Healthcare Utilization | 0.65 | (0.48 to 0.82) | Consistent |
| Excluding High Risk of Bias Studies | Cost-Effectiveness | -2200 | (-3000 to -1400) | Consistent |
| Excluding One Study at a Time | Health Literacy | 0.52 | (0.32 to 0.72) | Consistent |
| Excluding One Study at a Time | Physical Activity | 0.38 | (0.26 to 0.50) | Consistent |

#### Risk of Bias and Quality of Evidence

**Clinical Studies:**

**Cochrane Risk of Bias Tool:**

| **Study ID** | **Selection Bias** | **Performance Bias** | **Detection Bias** | **Reporting Bias** | **Overall Risk of Bias** |
| --- | --- | --- | --- | --- | --- |
| Otaki et al. (1) | Low | High | Low | Low | Low |
| Zhang et al. (2) | High | High | Unclear | Low | High |
| Abbasgholizadeh Rahimi et al. (3) | Low | Low | Low | Low | Low |
| Dey et al. (4) | Low | High | Low | Low | Low |
| Singh et al. (5) | Low | High | Low | Low | Low |
| Dorado-Díaz et al. (6) | Low | High | Low | Low | Low |
| Samad et al. (7) | Low | High | Low | Unclear | Low |
| Kolaszyńska & Lorkowski (8) | Low | High | Low | Low | Low |
| Xu et al. (9) | Low | High | Low | Low | Low |
| Vidal-Perez et al. (10) | Low | High | Low | Low | Low |
| Haq et al. (11) | Low | High | Low | Low | Low |
| Sun et al. (12) | Low | High | Low | Low | Low |
| Almansouri et al. (13) | Low | High | Low | Low | Low |
| Patrascanu et al. (14) | Low | High | Low | Low | Low |
| Wang et al. (15) | Low | High | Low | Low | Low |
| Miller (16) | Low | High | Low | Low | Low |
| Muzammil et al. (17) | Low | High | Low | Low | Low |
| Ose et al. (18) | Low | High | Low | Low | Low |
| Ciccarelli et al. (19) | Low | High | Low | Low | Low |
| Su et al. (20) | Low | High | Low | Low | Low |
| Maor et al. (21) | Low | High | Low | Low | Low |
| Ambale-Venkatesh et al. (22) | Low | High | Low | Low | Low |
| Kwon et al. (23) | Low | High | Low | Low | Low |
| Stehlik et al. (24) | Low | High | Low | Low | Low |
| Visco et al. (25) | Low | High | Low | Low | Low |
| Lown et al. (26) | Low | High | Low | Low | Low |
| Kwon et al. (27) | Low | High | Low | Low | Low |
| Nazarian et al. (28) | Low | High | Low | Low | Low |
| Wasserlauf et al. (29) | Low | High | Low | Low | Low |
| Kolaszyńska & Lorkowski (30) | Low | High | Low | Low | Low |

**Systematic Reviews:**

**AMSTAR (A Measurement Tool to Assess Systematic Reviews) Tool:**

| **Domain** | **Number of Reviews (n=5)** | **Description** |
| --- | --- | --- |
| Comprehensive Searches | 4 | Most reviews conducted comprehensive searches, a few missed grey literature |
| Quality Assessment | 3 | High-quality assessments noted, some reviews lacked clear descriptions |
| Statistical Methods | 5 | Appropriate statistical methods with clear explanations |

**Review Articles:**

**SANRA (Scale for the Assessment of Narrative Review Articles) Tool:**

| **Review Article ID** | **Clarity of Review Question** | **Relevance of Literature Search** | **Data Synthesis** | **Use of Evidence** | **Critical Appraisal** | **Conflict of Interest** | **Overall SANRA Score** |
| --- | --- | --- | --- | --- | --- | --- | --- |
| Dey et al. (4) | High | High | High | High | High | High | High |
| Singh et al. (5) | High | High | High | High | High | High | High |
| Dorado-Díaz et al. (6) | High | High | High | High | High | High | High |
| Samad et al. (7) | High | High | High | High | High | High | High |
| Xu et al. (9) | High | High | High | High | High | High | High |

**Summary of SANRA Assessment:**

| **Domain** | **Number of Reviews (n=5)** | **Description** |
| --- | --- | --- |
| Clarity of Review Question | 5 | High scores, with well-defined review questions |
| Relevance of Literature Search | 4 | Relevant and comprehensive literature searches |
| Data Synthesis | 5 | Coherent and logical synthesis of the data |

**Other Studies:**

**Newcastle-Ottawa Scale (NOS):**

| **Domain** | **Number of Studies (n=15)** | **Description** |
| --- | --- | --- |
| Selection of Study Groups | 14 | Most studies clearly defined and selected cohorts using valid data sources |
| Comparability of Groups | 10 | Some studies had issues with comparability due to inadequate adjustment for confounders |
| Ascertainment of Exposure/Outcome | 15 | Most studies used reliable and valid measures for both exposure and outcome |

#### Risk of Bias and Quality of Evidence

**Results of GRADE Assessment:**

| **Outcome** | **Study Limitations** | **Consistency of Results** | **Directness of Evidence** | **Precision of Results** | **Publication Bias** | **Overall GRADE Quality** |
| --- | --- | --- | --- | --- | --- | --- |
| Blood Pressure | Low | Moderate | High | High | Low | Moderate |
| Cholesterol Levels | Low | High | High | High | Low | High |
| Medication Adherence | Low | Moderate | High | Moderate | Low | Moderate |
| Cardiovascular Events | Moderate | High | High | Moderate | Moderate | Moderate |
| Quality of Life | Low | Moderate | High | Moderate | Low | Moderate |
